# A protein signature associated with active tuberculosis identified by plasma profiling and network-based analysis

**DOI:** 10.1101/2022.04.22.22274170

**Authors:** Zaynab Mousavian, Elin Folkesson, Gabrielle Fröberg, Fariba Foroogh, Margarida Correia-Neves, Judith Bruchfeld, Gunilla Källenius, Christopher Sundling

**Author notes:** corresponding author: Christopher Sundling. shared first author (Zaynab Mousavian, Elin Folkesson). shared last author (Judith Bruchfeld, Gunilla Källenius, Christopher Sundling).

## Abstract

**Objectives:** Tuberculosis (TB) is a bacterial infectious disease caused by *Mycobacterium tuberculosis*. Annually, an estimated 10 million people are diagnosed with active TB, and approximately 1.4 million dies of the disease. If left untreated, each person with active TB will infect 10 to 15 new individuals every year. Therefore, interrupting disease transmission by accurate early detection and diagnosis, paired with appropriate treatment is of major importance. In this study, we aimed to identify biomarkers associated with the development of active TB that can then be further developed for clinical testing.

**Methods:** We assessed the relative plasma concentration of 92 proteins associated with inflammation in individuals with active TB (n=19), latent TB (n=13), or healthy controls (n=10). We then constructed weighted protein co-expression networks to reveal correlations between protein expression profiles in all samples. After clustering the networks into four modules, we assessed their association with active TB.

**Results:** One module consisting of 16 proteins was highly associated with active TB. We used multiple independent transcriptomic datasets from studies investigating respiratory infections and non-TB diseases. We then identified and removed genes encoding proteins within the module that were low expressed in active TB or associated with non-TB diseases, resulting in a 12-protein plasma signature associated with active TB.

**Conclusion:** We identified a plasma protein signature that is highly enriched in patients with active TB but not in individuals with latent TB or healthy controls and that also had minimal cross-reactivity with common viral or bacterial lower respiratory tract infections.

## Introduction

Tuberculosis (TB), caused by *Mycobacterium tuberculosis* (Mtb), is an ongoing pandemic responsible for approximately 10 million clinical cases and 1.4 million deaths annually, making it the single deadliest infectious disease excluding the ongoing SARS-CoV2 pandemic. However, there are still many limitations in diagnostic methods available for active TB (1, 2). Around one-fourth to one-fifth of the world’s population is estimated to be latently infected with Mtb (3), of which 5-10% are estimated to eventually develop active disease.

Pulmonary TB is the most common clinical form, and it is diagnosed by detecting Mtb in sputum samples by microscopy, nucleic acid amplification tests, such as PCR, and the reference method mycobacterial culture. Several limitations to these methods exist; as compared with culture positivity, sputum microscopy is positive only in approximately 50% of cases (4) and PCR in approximately 90% of cases in respiratory samples (GeneXpert RIF/TB and GeneXpert Ultra) (5, 6). Additional difficulties relate to specific patient groups, such as HIV-infected patients and children, the latter commonly having low bacterial load and cannot produce sputum. Mycobacterial culture can take several weeks to yield positive results and requires specialized safety laboratories. In addition, it is often unavailable in resource-poor settings where TB is more prevalent. In addition, the diagnosis of extrapulmonary TB relies on invasive procedures to obtain samples for microbiological analysis. Thus, diagnosis is frequently based on clinical and radiological findings or algorithms, especially in low-resource settings. The diagnostic delays in endemic areas are well-described (7) The gap between estimated and reported TB cases was more than 4 million in 2020, and of those reported only 59% were microbiologically confirmed (8). No specific blood test or biochemical marker has yet been introduced in the routine clinical work-up to distinguish TB from other medical conditions. The need for non-sputum-based tests, both for screening and diagnostic purposes is urgent and the requirements of those tests have been described in detail in WHO statements for Target Product Profiles (9).

Over the last decades, various methods that examine the host response to Mtb infection have been evaluated. Attempts at identifying TB-specific transcriptional, protein, and metabolic signatures in patient blood samples were recently reviewed (10, 11). Promising results have emerged for transcriptional signatures (12, 13) but no protein signature for active TB has so far been validated in independent confirmatory studies. The protein signatures so far identified show limited overlap and together with varying study designs and methods, this makes a meta-analysis difficult (14).

In this study, we investigated the profile of 92 inflammatory proteins in the plasma from a Swedish cohort including individuals with active TB, latent TB, and healthy controls (Figure 1). Through weighted co-expression network analysis, we identified a signature that was highly associated with active TB and disease severity. We refined the signature by removing proteins associated with other bacterial and viral respiratory infections and sarcoidosis. We then validated the signature in several independent transcriptional datasets and showed it to be highly enriched in individuals with active TB.

**Figure 1:**
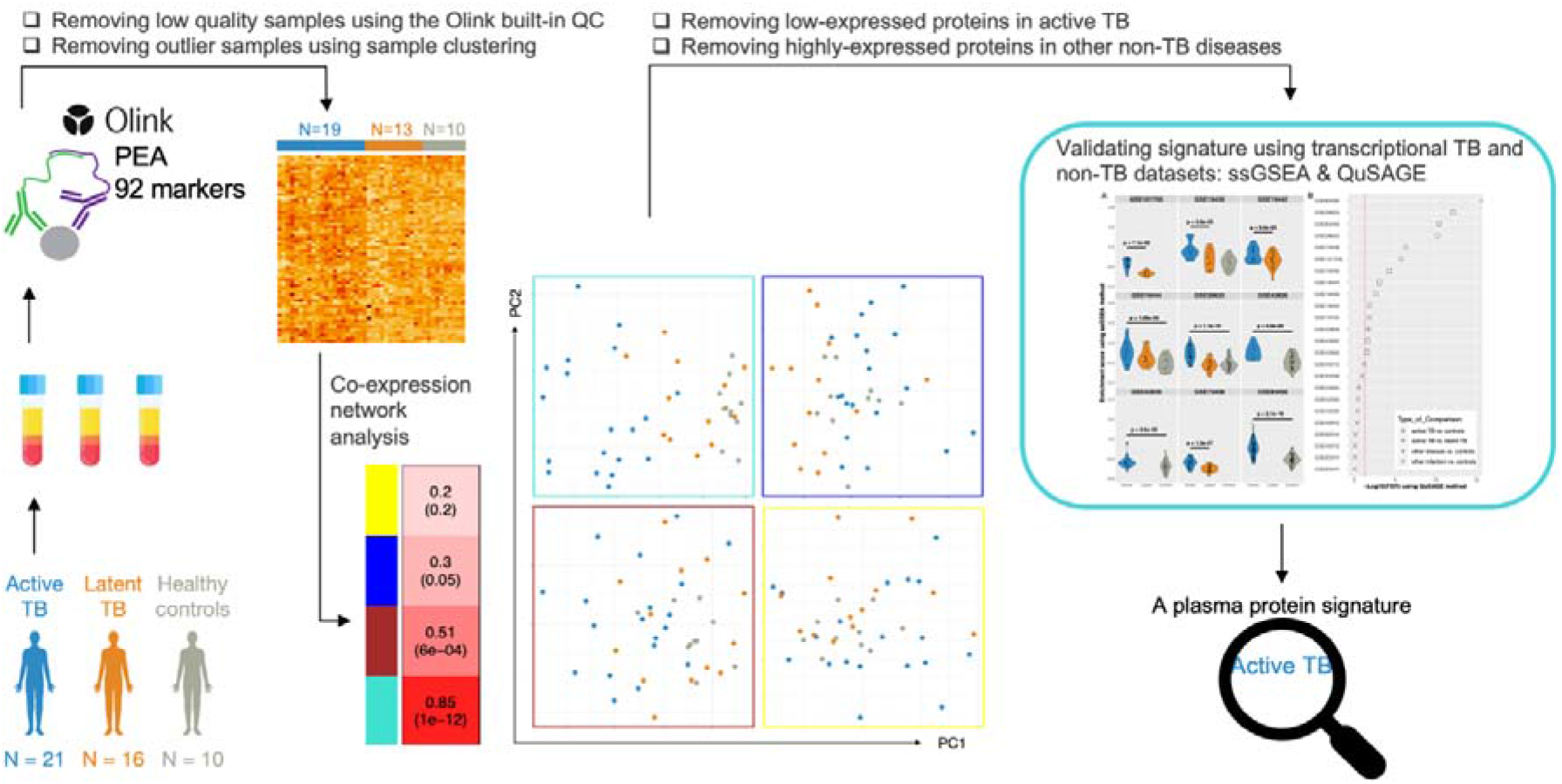
Flowchart of the study.

## Materials and methods

### Ethical considerations

The study was registered and granted ethical permission from the Swedish Ethical Review Board, EPN-number 2013/1347-31/2 and 2017/2262-32. All study subjects signed written informed consent forms after receiving written and verbal information, in relevant cases using professional interpreter services.

### Study participants

Patients were recruited at the Karolinska University Hospital, Stockholm from May 2018 to November 2019. Eligible for the study were: 1) Active TB patients within seven days of treatment initiation. Active TB was defined through microbiological verification via Mtb culture, or if culture negative, other microbiological positive result for Mtb (microscopy or PCR) combined with clinical and radiological findings and response to anti-TB treatment (active TB individuals are further described in Table S1); 2) Latent TB individuals with a positive IGRA result (QuantiFERON-TB Gold In-Tube (QFT-GIT) or QuantiFERON-TB Gold Plus (QFT-Plus) identified through contact investigation or screening of migrants, for which active TB had been excluded. The cut-off between recent and remote TB was set to two years after exposure; 3) Healthy controls with a negative IGRA-result and without previously known TB-exposure, co-morbidities, or immunosuppression.

### Data collection

Demographic, epidemiological, and clinical data for patients with active and latent TB were extracted from patient charts. For all subjects this included information regarding previous exposure to patients with active TB or infection, comorbidities and current medication, radiological, biochemical, and microbiological test results. For the active TB cases, clinical symptoms of TB disease were noted, and patients were classified according to pulmonary or extrapulmonary TB. Microbiological samples for mycobacterial analysis were collected independent of the study in accordance with clinical practice.

### Analysis of plasma proteins by proximity extension assay

Venous blood samples were collected in heparin vacutainer tubes and transferred to the laboratory for immediate preparation. The tubes were centrifuged at 670 *g* for 8 min after which the plasma fraction was aliquoted and stored at –80 °C.

Patient plasma was analysed using the Olink proximity extension assay, a qPCR technology that simultaneously measures combinations of cytokines in preselected panels. We used the 96-protein inflammatory panel, analysing 92 protein biomarkers and 4 controls (Table S2). In this automated process, specific antibodies carrying single stranded matching DNA bind in pairs to each of the target proteins allowing for DNA hybridization and subsequent DNA-extension. The resulting DNA, unique for each target protein, is then subject to PCR amplification and finally detection. The generated results consist of normalized protein expression (NPX) values that correspond to log2 transformed relative protein concentrations. The assay was performed by the Translational Plasma Profile Facility at SciLifeLab, Stockholm, Sweden.

### Protein expression data analysis

Repeated samples were used between experimental batches for running bridge normalization. We used *read_NPX* and *olink_normalization* functions from the OlinkAnalyze R package (https://github.com/Olink-Proteomics/OlinkRPackage) to read the log2 NPX protein expression values and perform bridge normalization between batches of our data respectively. Proteins with NPX values below the limit of detection (LOD) in more than 30% of samples were filtered out. To remove batch effects from the final dataset, we applied the *removeBatchEffect* function from the Limma R package (15) (Figure S1). We also performed differential expression analysis using the Limma R package.

### Weighted co-expression network analysis

We used the WGCNA R package to construct a weighted protein co-expression network among the proteins of our dataset. *Pearson correlation* and the *signedhybrid* network type were used in the *adjacency* function of the WGCNA package. In the *signedhybrid* network type, only links associated to positive correlations were retained in the network, and negative correlations were discarded. Since most of the biological networks have a scale-free topology, the WGCNA package in the *pickSoftThreshold* function tries to find the best value of the power parameter to make a scale-free network. After network construction, the network was clustered into modules containing proteins that were highly positively co-expressed using a hierarchical clustering algorithm as implemented in the *cutreeDynamic* function of the WGCNA package with parameters deepSplit = 4 and minClusterSize = 5, and other parameters set as default. The expression profiles of proteins in each module were summarized by module eigengenes. In the *moduleEigengenes* function from the WGCNA package, the first principal component of the expression data of each module is measured as a module eigengene for that module. To find a module associated with TB progression, the correlation between each module eigengene and the trait vector was computed to identify which module that had a significant correlation with TB progression.

### Protein signature enrichment analysis

To further validate the protein signature obtained from the co-expression network analysis, we applied two computational methods (ssGSEA and QuSAGE) for enrichment analysis in independent transcriptomic datasets. In ssGSEA, the enrichment scores of the protein signature were calculated per sample based on the absolute value of proteins expression in that sample to quantify how much the protein signature was overrepresented in a specific sample. Moreover, to verify that the protein signature was specific to active TB and not to other non-TB respiratory infections or diseases with clinical presentations similar to active TB, the *qusage* function from the QuSAGE R package was applied to datasets comparing active TB or non-TB disease to various control groups. The gene set differential expression was calculated by combining individual probability functions obtained from individual differential expression of genes in a particular comparison. A p-value, calculated by QuSAGE, determined the statistical significance of a gene set for each given comparison. We also used different types of single sample gene set enrichment analysis algorithms, including ssGSEA, GSVA (16) and zscore (17), all implemented in the gsva R package, to compare all co-expressed modules in terms of enrichment in active TB versus latent TB and healthy controls.

## Results

### Clinical characteristics of the study participants

The plasma of 21 individuals with active TB, 16 individuals with latent TB, and 10 healthy controls were analyzed using the Olink inflammation proximity extension assay. During data processing, 5 samples (2 active TB and 3 latent TB) were excluded for either failing the quality control check included in the OlinkRPackage or identified as outliers by cluster analysis (Figure S2). The characteristics of the remaining study participants are further described in Table 1.

**Table 1.** Clinical characteristics of the sample cohort

|  |  | Active TB<br>(n=19) | Latent TB<br>(n=13) | Healthy<br>controls<br>(n=10) |
| --- | --- | --- | --- | --- |
| Men n (%) |  | 8 (42) | 8 (62) | 6 (67) |
| Age, y (mean, range) |  | 39 (20-71) | 39 (18-72) | 25 (21-37) |
| Origin from TB high endemic country, n |  | 14 | 5 | 0 |
| Time since immigration to Sweden, y |  | 9 (0.2-45) | 5 (0.5-15) | n/a |
| BCG-vaccination | yes/no/unknown | 2/1/16 | 4/1/8 | 1/9/0 |
| IGRA result | pos/neg/unknown | 10/1/8 | 13/0/0 | 1<sup>6</sup>/9/- |
| Previous Mtb<br>infection | active<sup>1</sup> | 3 | 0 |  |
|  | latent | 1 | 0 |  |
| Previous Mtb<br>exposure<sup>4</sup> | <2 years (recent) | 4 | 10 |  |
|  | >2 years (remote) | 1 | 1 |  |
| **Biochemistry**<sup>2,5</sup><br>(mean, range) | CRP (mg/L) | 25 (1-94) | 1 (1-2) |  |
|  | ESR (mm) | 51 (7-117) | 6 (1-21) |  |
|  | WBC (10<sup>9</sup>/L) | 6.5 (3.8-11.8) | 6.6 (5.6-9) |  |
|  | Hb (g/L) | 126 (102-149) | 145 (118-158) |  |
|  | Alb<sup>3</sup> (g/L) | 32 (26-38) | 41 (39-44) |  |

Patient origins; Active TB: Somalia (4) Eritrea (3), Sweden (3), Philippines (2), Peru, Bangladesh, Pakistan, Mongolia, Poland, China, Afghanistan. Latent TB: Sweden (3), Romania (3), Moldavia (2), Mongolia, Ethiopia, Eritrea, Ghana, India.
Comorbidities; Active TB: postpartum period, hypertension (2), intestinal schistosomiasis, chronic hepatitis B, acute thyroiditis. Latent TB: chronic hepatitis B, hypertension (2)
C-reactive protein (CRP), Erythrocyte sedimentation rate (ESR), White blood cell count (WBC), Hemoglobin (Hb), Albumin (Alb)
<sup>1</sup>Two patients previously treated for ATB, 1 and 4 years before; one patient not treated
<sup>2</sup>ATB n=19 LTBI n=9
<sup>3</sup>ATB n=17 LTBI n=7
<sup>4</sup>Exposure self-reported or verified
<sup>5</sup>Normal range; CRP<5, ESR <20, WBC 4.4-10.0, Hb >120 (F), >130 (M), Alb>38
<sup>6</sup>Healthy controls; one QFT borderline (0.36/0.39), no exposure to TB

As a reflection of the low TB incidence in Sweden, most study individuals originated from other countries, mainly situated in Africa, Asia and Eastern Europe. Fifteen of 19 active cases had pulmonary TB including 3 with pleuritis, 6 of which were sputum smear positive. Four had extrapulmonary TB cases of which two with disseminated disease. Only 1 out of the 19 active TB patients was not confirmed by microbiologic culture; this patient had a positive PCR for *M. tuberculosis* in a lymph node aspirate and gastric lavage as well as radiologic signs of active pulmonary TB and showed a clinical response to TB-treatment. The active TB cases were sampled within one week of treatment initiation except for one patient that was sampled at day 9. Of the 13 latent TB patients, 10 had a known recent TB exposure 1-4 months prior to study inclusion. All but 2 latent TB individuals completed preventive TB treatment, and none progressed to active TB during a follow up period of > 2 years. There were few significant co-morbidities, with no patient in either group on immunosuppressive treatment. All active TB patients were HIV negative. The latent TB and healthy controls were not routinely tested for HIV infection.

From the 10 healthy controls, 1 had a QFT-Gold Plus result in the low range of the borderline interval (0.35-0.99 IU/ml) but with no known exposure, potentially indicating a false positive result (18, 19).

### Network construction reveals one module associated with active TB

Following data pre-processing, including quality control, batch correction, and clustering the relative level (NPX) of each of the 92 proteins was assessed. Proteins with more than 30% of samples with NPX values below the limit of detection (n = 28) were excluded from further analysis. Out of those 28 proteins, 25 had no differential expression between the groups (Figure S3). A weighted protein co-expression network was constructed with the remaining 64 proteins to examine correlations between protein expression profiles in all included samples (n = 42 individuals). We considered only the links associated with positive correlations in the network reconstruction and selected power 8 to reach a scale-free topology (Figure S4). After clustering the network into modules, four modules indicated by distinct colours in Figure 2 were discovered (Table 2), including 34 proteins. The remaining 30 proteins that were not included in any of the modules, were discarded from further analysis. We observed that the turquoise module out of the four modules had a significantly stronger correlation with active TB (Table 2). A visual representation of the protein co-expression network and the discovered protein modules were then generated using Cytoscape 3.0 (20) (Figure 2).

**Table 2:**
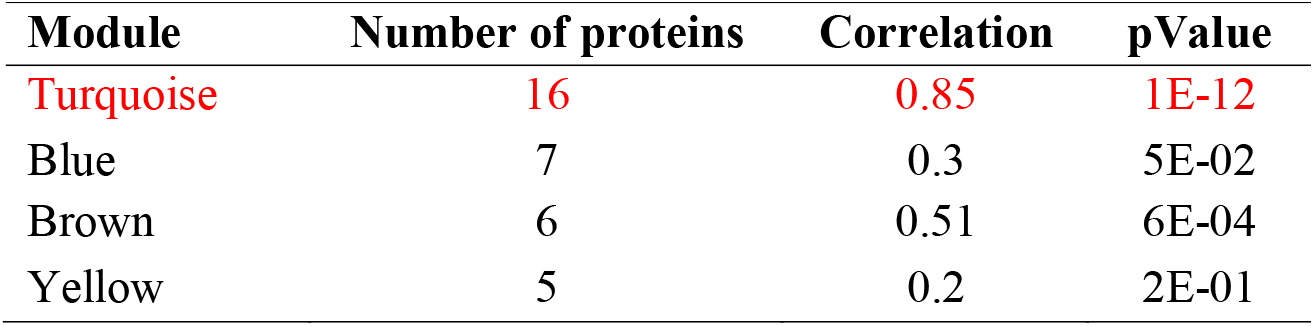
Details about the modules of the protein co-expression network

**Figure 2.**
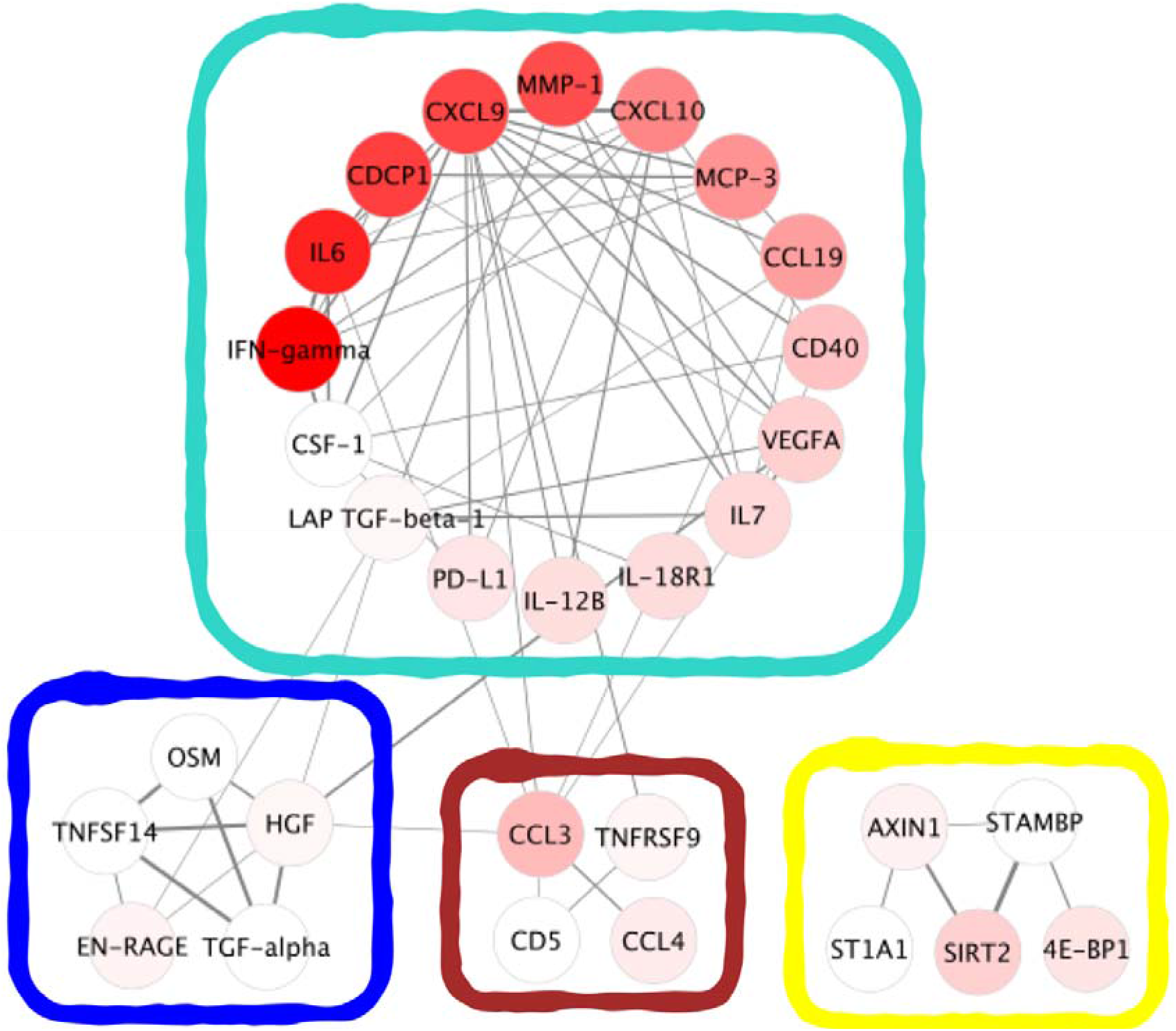
A visual representation of the protein co-expression network and the protein modules: The protein modules are surrounded by the colours turquoise, blue, brown or yellow. Nodes are coloured based on protein abundance (Fold Change) in comparison of active TB vs. control, with a darker red indicating a larger fold change. Links are depicted with thickness proportional to the correlation and only links representing Pearson’s r > 0.6 are shown.

We then assessed how the markers differed between individuals with active TB, latent TB, and healthy controls (Figure S5). All differentially expressed proteins (log2(FC) ≥ 1 and FDR-corrected p-value < 0.01) between either active TB and latent TB or active TB and healthy controls except FGF-21 and EN-RANGE were included in the turquoise module (Figure 3). Eight of the 16 proteins in the turquoise module were highly expressed in active TB patients while the remaining 8 proteins were strongly co-expressed with those highly expressed proteins (Figure 2). To illustrate how the turquoise module segregated active TB individuals from latent TB and healthy controls, we performed principal component analysis (PCA) on the expression profile of proteins of each module and used the first and the second principal components (PC1 and PC2) to show differences in protein levels (Figure S6). All active TB samples except one were clearly separated from the other samples by the proteins in the turquoise module, suggesting that the included proteins can potentially serve as a signature to identify individuals with active TB.

**Figure 3.**
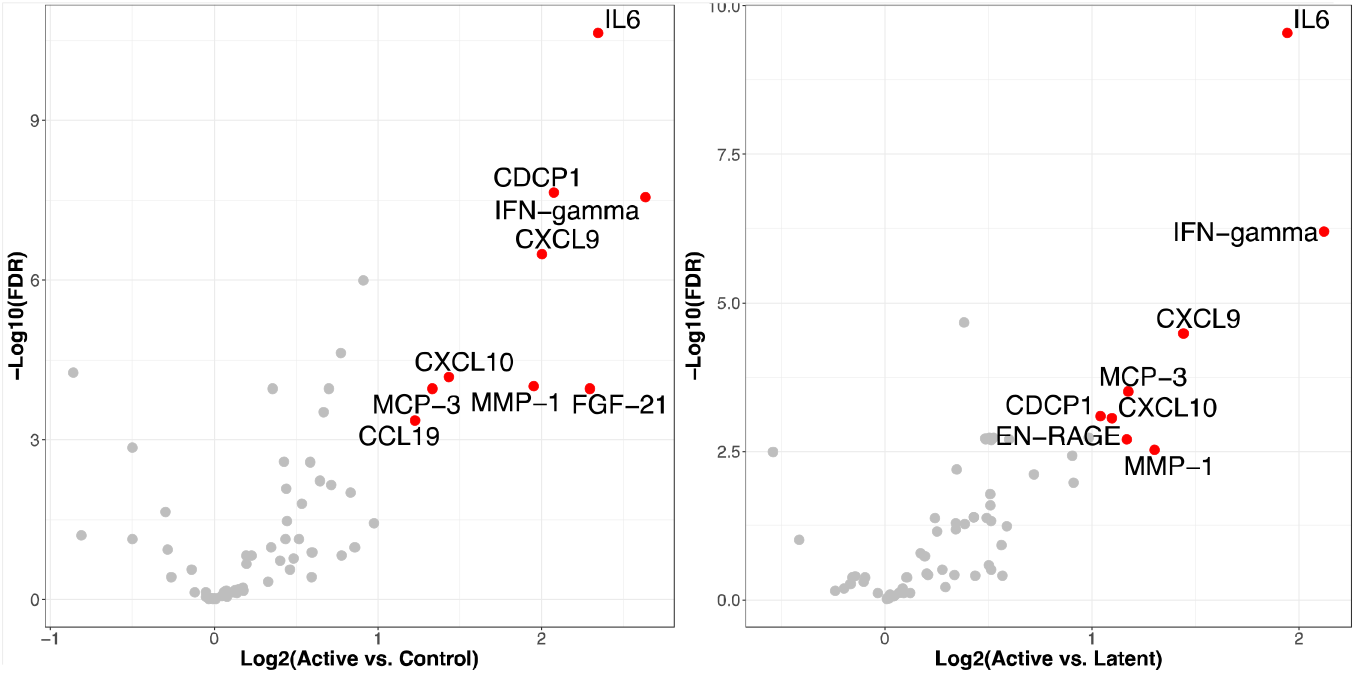
Differential expression analysis of all 64 proteins: Active TB vs. control (left) and active TB vs. latent TB (right). Red points indicate the significant highly expressed proteins with log2(FC) > 1 and FDR-corrected P-value < 0.01 in each comparison.

### Validating the turquoise module in independent transcriptomic TB datasets

There is limited overlap in markers between studies investigating protein or gene signatures aiming to discriminate active TB from latent TB, healthy controls, or other diseases (Figure S7). Additionally, we did not identify any available pre-processed proteomic dataset including all proteins of the turquoise module that we could use for validation. For this reason, we chose to instead evaluate to what extent the proteins of the different modules could distinguish active TB from latent TB and healthy controls in transcriptomic datasets. We applied all four modules to several independent cohorts using multiple gene set enrichment analysis methods. Nine transcriptomic TB datasets were selected based on the criteria of age >15, number of individuals per group >10 and no anti-TB treatment (Table S3). The datasets were provided in the curatedTBData R package (21), which also included the corresponding genes to all proteins included in the four modules. In total the datasets we selected included >3000 individuals from four continents. Three enrichment methods (ssgsea, gsva and zscore) demonstrated high enrichment of the turquoise module in active TB and low enrichment in latent TB and the control group across most of the TB datasets (Figures S8-S10). This was also the case for the blue module, although to a less extent compared with the turquoise module. In contrast, neither the brown nor yellow modules were found highly enriched in active TB using the same three methods on the same transcriptomic datasets (Figures S8-S10).

### Identification of a new 12-protein signature for active TB diagnosis

We identified the turquoise module as having the highest correlation with active TB. However, two of the 16 proteins in the module, LAP TGF-beta-1 and CSF-1, had low fold change values (log2(FC) < 0.5) when comparing active TB versus both latent TB and healthy controls (Figure 3), and were therefore potentially redundant.

To investigate if the remaining 14 proteins were specific for active TB, we performed differential expression analysis using multiple transcriptomic datasets including either viral or bacterial lower respiratory tract infections (LRTI), systemic inflammatory response syndrome (SIRS) or sarcoidosis (Figure 4). This allowed us to identify genes encoding the signature proteins that were highly and significantly expressed (log2(FC) > 1 and P-value < 0.05) in diseases with clinical symptoms overlapping with active TB. To this end, we used three transcriptomic datasets, GSE42026 (22), GSE40012 (23) and GSE60244 (24), each containing various types of lower respiratory infections. GSE42026 included (pediatric) patients with severe LRTI of different etiologies; bacterial, (mostly *Streptococcus pneumoniae*), Influenza A/H1N1/09 and Respiratory syncytial virus (RSV) infection. GSE40012 included adult patients with severe community acquired pneumonia (CAP); either bacterial or caused by Influenza A/H1N1, and SIRS (25), without evidence of infection. Finally, GSE60244 included patients hospitalized for bacterial LRTI (*Streptococcus pneumoniae* being most common), viral LRTI (Influenza A, B or RSV) and viral/bacterial coinfection. We also conducted the same experiment using three datasets from two studies containing sarcoidosis samples (26, 27). Two genes, corresponding to the proteins IL18R1 and CXCL10 (IP-10), stood out in these analyses (Figure 4). IL18R1 was highly expressed in severe viral and bacterial LRTI and SIR, while CXCL10 was highly expressed in severe viral LRTI and in one of the sarcoidosis studies. However, most of the proteins of the signature were only observed to be expressed at high levels in individuals with active TB. Therefore, in addition to LAP TGF-beta-1 and CSF-1 that were expressed only at very low levels, we also removed IL18R1 and CXCL10 from the signature, leading to a 12-marker plasma signature associated with active TB and with low cross-reactivity to other bacterial/viral lower respiratory infections and sarcoidosis. The final signature consisted of the proteins IFN-gamma, IL6, CDCP1, CXCL9, MMP-1, MCP-3, CCL19, CD40, VEGFA, IL7, IL-12B and PDL-1.

**Figure 4.**
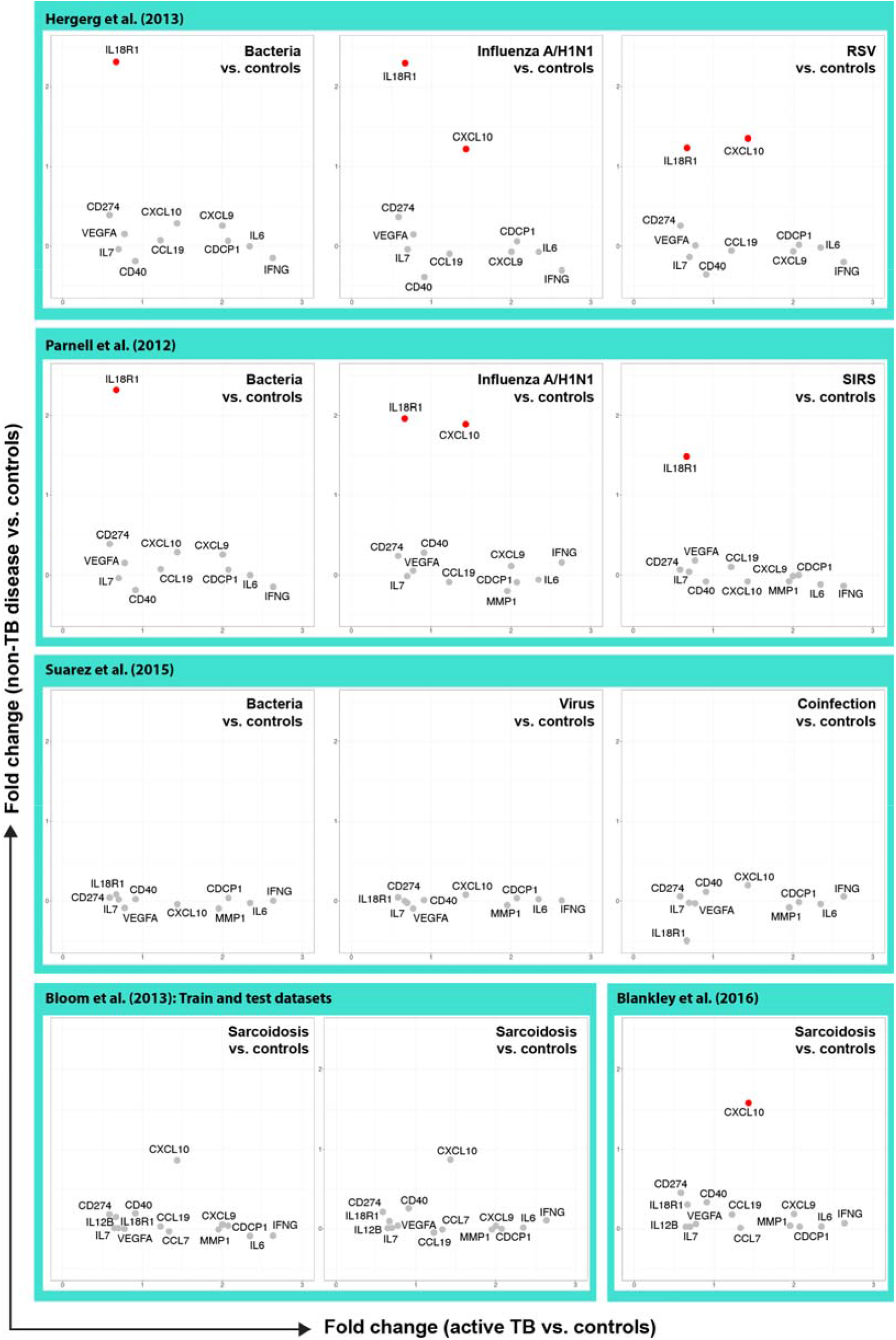
The expression of proteins of the turquoise module in our Swedish cohort compared to other viral/bacterial infections, SIRS and sarcoidosis from different datasets. The fold-changes of proteins of the turquoise module in our cohort (active TB vs. controls; x-axis) compared to their fold-changes in other non-TB diseases (non-TB diseases vs. controls; y-axis). Red points indicate proteins of the turquoise module, which genes encoding them are significantly highly expressed in either other infections or other diseases (log2(FC) > 1 and P-value < 0.05). Row 1). Pediatric patients with severe respiratory tract infection: Bacterial: (n=18, *Streptococcus pneumonia* (12), *Streptococcus pyogenes* (4) *Staphylococcus aureus* (2) including 5 with viral co-infection (non-Influenza A H1N1/RSV); Influenza A/H1N1 (n=19), RSV (n=22) HC (n=33). Row 2) Adult patients with severe CAP or SIRS requiring ICU-care: Bacterial: (n=16, mixed etiology) Influenza A/H1N1(n=8), SIRS without infection (n=12), HC (n=18). Row 3) Adult patients hospitalized for LRTI: Bacterial (n=22; *S. pneumoniae* (13), *Moraxella catarrhalis* (4), *S. aureus* (2), mixed bacterial (3); viral (n=71, Influenza A (32), RSV (17), Influenza B (9), HMPV (7); mixed bacterial/viral (n=25) HC (n=40). Row 4) pulmonary sarcoidosis vs controls. RSV = respiratory syncytial virus; CAP = Community acquired pneumonia; ICU = Intensive Care Unit; SIRS = systemic inflammatory response syndrome; LRTI = Lower respiratory tract infection; HMPV = human metapneumovirus.

### Validating the 12-marker signature in TB and non-TB proteomic and transcriptomic datasets

To assess the significance of the 12-marker signature in independent TB cohorts, we performed ssGSEA on transcriptomic TB datasets from the curatedTBData R package (Table S3). We observed significantly (p < 0.05) higher enrichment scores in the active TB group, compared with either latent TB or controls in all the transcriptomic TB datasets except one (GSE19444) (Figure 5A). We then used QuSAGE to compare the enrichment of the 12-marker signature in TB infections with both other bacterial/viral infections and other pulmonary diseases (Figure 5B). Three different comparisons were done using the GSE42026, GSE40012 and GSE60244 datasets to assess the signature in respiratory infections versus healthy controls, and in sarcoidosis disease versus healthy controls using three transcriptomic datasets (GSE83456, GSE42826 and GSE42830) (Figure 5B). We observed that the 12-marker signature was significantly overrepresented (FDR corrected P-value < 0.05) in all comparisons between active TB and latent TB or healthy controls but was not significantly enriched in those gene sets comparing other LRTIs vs. healthy controls. However, in two of the three sarcoidosis datasets the signature was also enriched, indicating that the signature might not, on its own, be able to distinguish active TB from sarcoidosis without also weighing clinical data.

**Figure 5.**
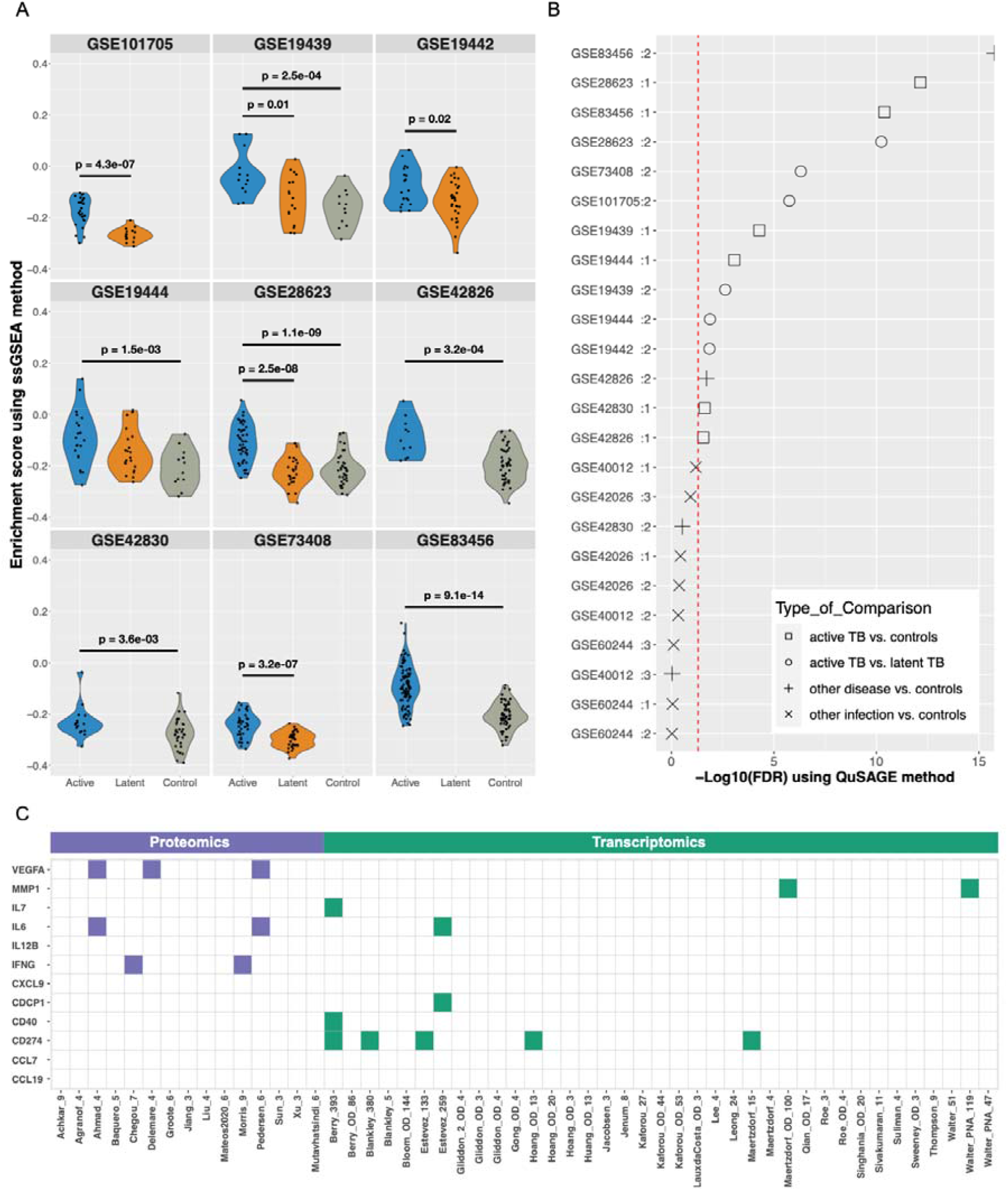
Validation and overlap of the 12-marker protein signature. A) ssGSEA indicating enrichment score for the 12-marker signature in different transcriptomic datasets. Blue indicates individuals with active TB, orange indicates latent TB and grey indicates healthy controls. The p values show the statistical significance for the enrichment score difference between either active TB and Latent TB or Active TB and healthy controls using the wilcox.test. B) QuSAGE analysis with -log(FDR) indicating capacity of the 12-marker signature in separating active TB from healthy controls (open boxes), active TB from latent TB (open circles), non-infectious inflammatory disease from healthy controls (+) and other LRTIs or SIRS from healthy controls (×). The red line indicates FDR<0.05. C) Overlap between individual proteins in the 12-marker signature with other proteomic signatures (left) and transcriptomic signatures (right) from the TBSignatureProfiler R package identifying active TB.

We then compared the 12-marker signature with the other published gene signatures from the TBSignatureProfiler R package (28) and published protein signatures to investigate overlap between protein and transcriptional signatures with the proteins of our signature (Figure 5C). VEGFA, IL6, and IFN-gamma were identified in at least two other proteomic studies (29-33), while CD274 (also called PD-L1) was observed in several published transcriptional signatures. IP-10 (also called CXCL10), which was removed due to its high expression in severe viral LRT and sarcoidosis appears in several proteomic studies (30-33). The other markers were less common or identified in the current study. Although these proteins have not been included in protein signatures before, they have been associated with TB (34-37) and could potentially be generated via similar signaling pathways in response to mycobacterial infection, such as has been indicated for signal transducer and activator of transcription 1 (STAT1) in TB (38). To assess if this was the case, we used the StrongestPath application (39) in Cytoscape to evaluate how the proteins were connected to different STAT transcription factors based on data from the KEGG database (Figure S11). We observed that several of the proteins were directly associated with STAT1, consistent with previous literature (38) to a lesser extent with STAT3 and STAT4, and indirectly with STAT2 and STAT5A.

### Association between the 12-marker signature and disease severity

Principal component analysis of the 12-marker signature (Figure 6A) showed that, similar to the turquoise module (Figure S6), active TB was primarily separated from latent TB and healthy controls by principal component 1 (PC1). We therefore hypothesized that PC1 could represent a proxy for the magnitude of the inflammatory response and disease severity. To evaluate if this was the case, we stratified the individuals with active TB based on their PC1 values and a set of clinical features (Figure 6B). The heatmap shows that samples with a smaller PC1 value, located towards the left side of heatmap, were associated with several biochemical markers indicating more extensive inflammation, such as lower hemoglobin (Hb) and albumin (Alb), and higher erythrocyte sedimentation rate (ESR) and C-reactive protein (CRP). The presence of any systemic symptoms (fever, night sweats and weight loss) and smear positivity were also associated with a lower PC1 value. We next performed a regression analysis to see if individual or combinations of the clinical variables could explain the PC1 values. A standard least squares model including ESR (estimate: –0.03, 95% CI: – 0.016 - –0.044, p = 0.0008) and Alb (estimate: 0.21, 95% CI: 0.08-0.34, p = 0.006) were highly associated with PC1 (r^2^ = 0.74, p < 0.0001), clearly associating the signature with clinical presentation (40).

**Figure 6.**
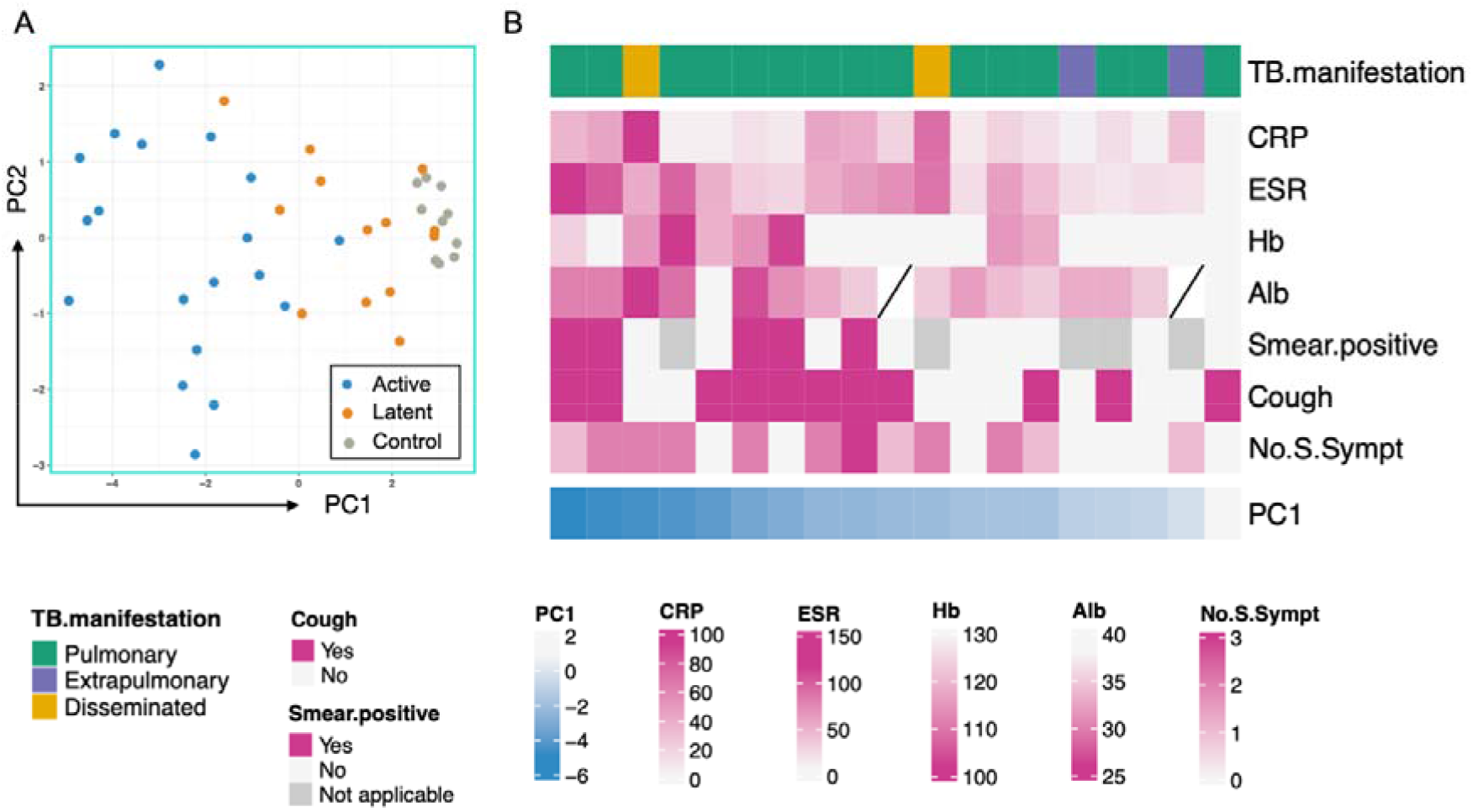
Association between the first principal component (PC1) and clinical characteristics of active TB. A) PC1 and PC2 indicate variation in concentrations of the 12-marker signature for individuals with active TB (blue dots), latent TB (orange dots) and healthy controls (grey dots). B) Clinical characteristics (rows) of all active TB donors (columns) stratified by the negative value of PC1 (the more to the left indicating a stronger signature). TB manifestation: Pulmonary: (including 3 with pleuritis); 2 disseminated (pulmonary/abdominal/lymphnode and liver/lymphnode) and 2 extrapulmonary (lymphnode and soft tissue/osteitis), CRP = C-reactive protein, ESR = erythrocyte sedimentation rate, Hb = hemoglobin, Alb = albumin. Smear.positive; sputum smear microscopy positive (5 of 14 with sputum); Cough: as reported by patient; Presence of systemic symptoms (fever, weight loss, night sweats); No.S.Sympt = number of systemic symptoms; night sweats, fever, and weight loss.

The three active TB cases with the weakest PC1 signal had few symptoms and limited disease activity and inflammation (Figure 6A). One patient had a culture-confirmed lymph node TB. Another patient had a PCR positive but culture-negative lymph node and pulmonary TB. The active TB case with the lowest inflammatory signal was found through contact investigation very early in active disease progression with cough as the only symptom but with no inflammation in laboratory tests. This suggests that the signature has the potential to identify active TB very early on in the progression from latent TB.

Although the signature was not optimized for the detection of latent TB, the visually apparent separation of latent TB from healthy controls (Figure 6A) could possibly indicate a TB-specific immune response detectable in the plasma of these individuals. Two of the three latent TB cases clustering with the healthy controls had no recent exposure to TB and were clinically classified as remotely acquired infections (i.e., no known exposure to a TB case for > 2 years). In summary, these data suggest that the signature could be helpful in identifying TB disease progression or cure.

## Discussion

The purpose of this study was to identify a protein signature in plasma that was enriched in individuals with active TB compared to those with latent infection and healthy controls. We used the Olink PEA method to simultaneously measure 92 proteins in plasma. Using co-expression analysis, we could identify 16 proteins that were co-expressed and highly correlated with active TB. To test the validity of these findings, we used publicly available transcriptomic datasets. First, we removed two proteins with very low differential expression between active TB and controls. We then refined the signature by excluding two additional proteins that were highly expressed in other lower respiratory tract infections. This allowed us to generate a 12-protein signature that was highly specific when applied to independent TB datasets. When retesting the signature on our own data it also retained a similar discriminatory capacity as the initial 16-protein signature.

Albeit small, the study cohort is well defined, with microbiological verification of all active TB cases, a wide range of disease severity and with large variation in patient origin, reflecting the TB population in Sweden and thus increasing the likelihood of generalizability to other geographic regions. Further, results were validated in nine transcriptomic data sets including over 3000 individuals from four continents.

On an individual patient level, the signature was associated with disease activity with a stronger protein signature significantly related to perturbations in common clinical inflammatory markers (ESR and albumin) and reported symptoms.

We have shown that the identified signature is differentially expressed in independent TB transcriptomic datasets and importantly, it is not expressed in patients with other lower respiratory tract infections, which is essential if the signature is to be used as a diagnostic screening test in a clinical setting as intended. However, when testing the signature in a sarcoidosis dataset (27), we observed a significant enrichment of the proteins. Sarcoidosis and TB have previously been shown to have similar gene expression patterns (27). Both diseases demonstrate an interferon-driven gene up-regulation, although as shown by Blankley et al, on a group level this pattern is more strongly expressed in active TB, reflecting disease activity (27). We were not able to directly compare sarcoidosis versus active TB to assess if the signal is higher in one group or the other. However, if the signature was to be used as a screening test this potential overlap will likely not pose a significant diagnostic problem in a clinical setting as symptoms and other clinical information such as radiology will help separate the two conditions. Further, in TB high-endemic countries where a screening test is most urgently needed, the prevalence of sarcoidosis is very low compared to pulmonary TB (41, 42).

Lack of overlap between protein signatures for active TB has been previously described, and the methods used to quantify proteins in different studies also vary (14). When comparing the 12-marker signature to recently published protein signatures (29-33, 40, 43-51) some biomarkers reappear, although the overlap is limited. Additionally, in the published proteomic studies there were none with processed data that included all the proteins of our signature in their dataset, making us unable to use them to validate the signature proposed in this study.

VEGF and IL-6, present in the 12-protein signature, together with IL-8 and IL-18, constitute a 4-protein signature identified by Ahmad et al. (29). They analyzed 47 proteins with Luminex and the 4 selected proteins were validated in serum from three different patient cohorts collected through the FIND initiative. The sensitivity for active TB in TB suspects was 80% (95% CI, 73 to 85%), and the specificity was 65% (95% CI, 57-71%). Interestingly, there was quite a large overlap between the Olink inflammation panel we used and their 47-protein Luminex panel. Although IL-8 and IL-18 were more abundant in active TB patients compared with healthy controls in our dataset (Figure S5), the fold-change values were low compared with the proteins included in the 12-protein signature and were not significantly higher when compared with latent TB individuals.

VEGF was also included in a 4-protein signature (CCL1, CXCL10, ADA-2, VEGF) proposed by Delemarre et al. (30), where they compared active TB to treated and untreated latent TB. The signature was validated in two separate patient cohorts with a sensitivity of 95% and a specificity of 90%. CXCL10 was also highly expressed in our analysis but was removed due to its high expression in other lower respiratory tract infections. CCL1 and ADA-2 were not analyzed in the current study.

Although VEGF was not included in the Chegou et al. (31) 7-protein signature, it was increased in active TB patients in their cohort. Of their 7 proteins, IFN-gamma overlaps with our 12-protein signature. CXCL10 (IP-10) again appears in their signature while the other proteins (CRP, TTR, CFH, APO-A1, SSA) were not part of the proteins evaluated in our study.

In 2021, Morris et al. (32) and Mutavhatsindi et al. (51) investigated the same 22 proteins as Chegou et al. (31) and attempted to validate the 7-protein signature in patients with suspected TB. In the first study, the sensitivity was very high (98%) while the specificity was low (12%). They argued that this might be due to the different patient cohorts – primary care level versus hospital care level. Instead, they identified a 9-protein signature (fibrinogen, alpha-2-macroglobulin, CRP, MMP-9, transthyretin, complement factor H [CFH], IFN-gamma, IP-10, and TNF-alpha) where IFN-gamma and IP-10 reappear. In the test set, the sensitivity was 92% (95% CI, 80-98%) and specificity 71% (95% CI, 56-84%) for diagnosing culture verified TB from other diseases in TB suspects. Their second study (51) failed to validate the 7 marker signature. Instead, they proposed CRP and CCL1 as a signature that performed equally well in both HIV– and HIV+ individuals. Trying to design a protein signature based on previous findings, Pedersen et al. (33) evaluated 9 proteins in pulmonary TB patients compared to healthy controls. They found that IL-6, VEGFA, (and IP-10) are significantly increased in active TB. Their proposed signature consists of IP-10 and 4 miRNA molecules, including miR-29a, miR-146a, miR-99b and miR-221. IP-10 is also part of the 5-protein signature identified by Luo et al. (52).

Garay-Baquero et al., analyzed more than 5000 peptides using Mass spectrometry on a relatively small discovery cohort (10 individuals with active TB and 10 healthy controls) (44). They identified 46 proteins to be overexpressed in active TB and selected 9 and 7 proteins for validation in larger populations in South Africa and the UK, respectively. They used Luminex or ELISA and compared active TB to other respiratory diseases (ORD) and healthy controls. The proposed 5 protein signature; CFHR5, LRG1, CRP, LBP, and SAA1, performed well with an AUC > 0.8 in both settings for active TB vs ORD. There was no overlap with the 12-protein signature identified here, and none of the proteins were among the 136 proteins identified as associated with active TB (46 more abundant and 90 less abundant) in their discovery phase, although the use of different methods makes direct comparison difficult.

Another large proteomic study by de Groote et al., (53) used SOMAscan to measure over 4000 proteins in 1470 patient samples from pulmonary TB patients and other TB suspect cases and resulted in a 6-protein signature. Although their signature did not overlap with our 12-protein signature, IL-6 and MMP-1 were overexpressed in their active TB cases.

In summary, IFN-gamma, IL-6, and VEGF, together with IP-10 were identified as markers for active TB in several previous studies, as well as identified in our study. These are all regulated by STAT-1, which has previously been identified as an important immune response pathway in Mtb infection (38), and potentially explains why these proteins are identified in different studies. However, as found here, IP-10 is also highly expressed in other severe infectious diseases and as such is likely not specific for TB in the absence of Mtb-specific stimulation (54).

To conclude, we identified a plasma biomarker signature associated with active TB progression that was further corroborated in several independent datasets. Although the signature can likely be optimized further by testing it in independent protein datasets, the included biomarkers warrant further investigation and development for diagnostic purposes, which will be critical to stopping the TB pandemic.

## Data Availability

All data produced in the present study are available upon reasonable request to the authors

## Acknowledgements

We would foremost like to thank the study participants for agreeing to be included in the study and donate study material. We would also like to thank the study nurses Monica Modin, Jan Bellbrant, Anna Dahlberg and Lena Jansson for patient inclusion, Julius Lautenbach and Victor Yman for constructive discussions about the analysis, and the SciLifeLab Translational Plasma Profiling facility for their support and generating data for this study. This study was supported by grants from the Swedish Research Council (2021-03706), Magnus Bergvall Foundation (2019-03436), Åke Wiberg Foundation (M19-0559) and the Swedish Medical Association (SLS-934363) to CS and grants from Swedish Research Council (2019-04663 and 2020-03602) and the Heart- and Lung Foundation (20180386 and 20200194) to GK.

