## Supplemental material for "A protein signature associated with active tuberculosis identified by plasma profiling and network-based analysis"

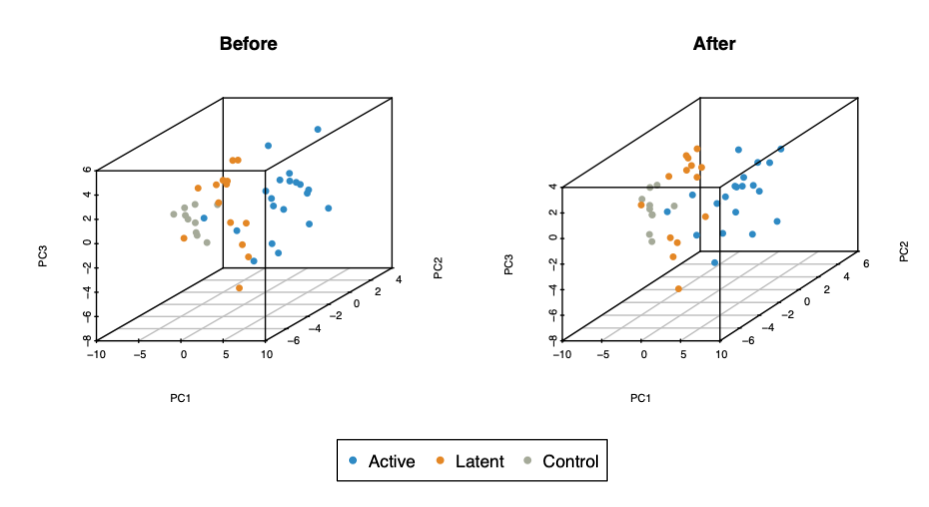

Figure 1. Visualizing samples of different groups: before and after batch effect removal.

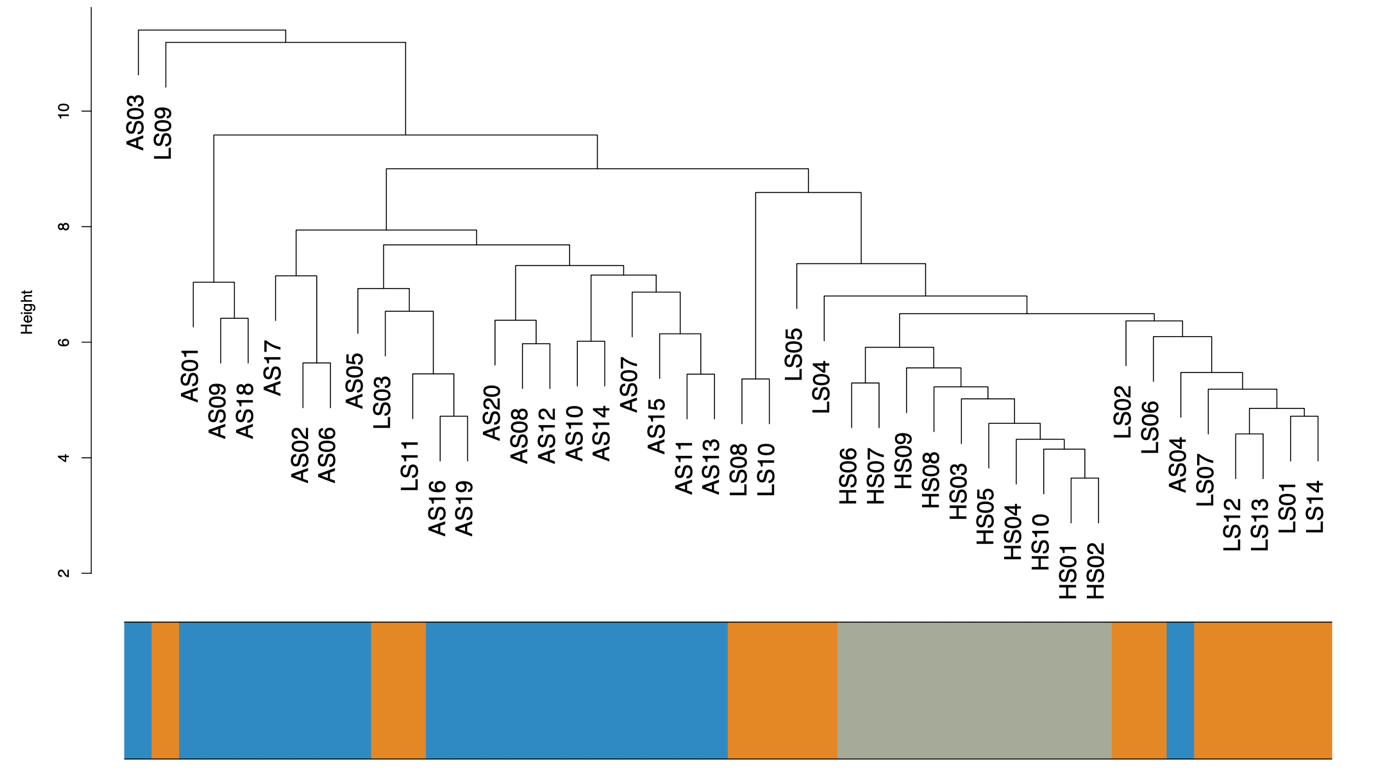

Figure 2. Hierarchical samples clustering to detect outliers. Two samples, one from active TB group and one from latent TB group, were identified as outliers and removed.

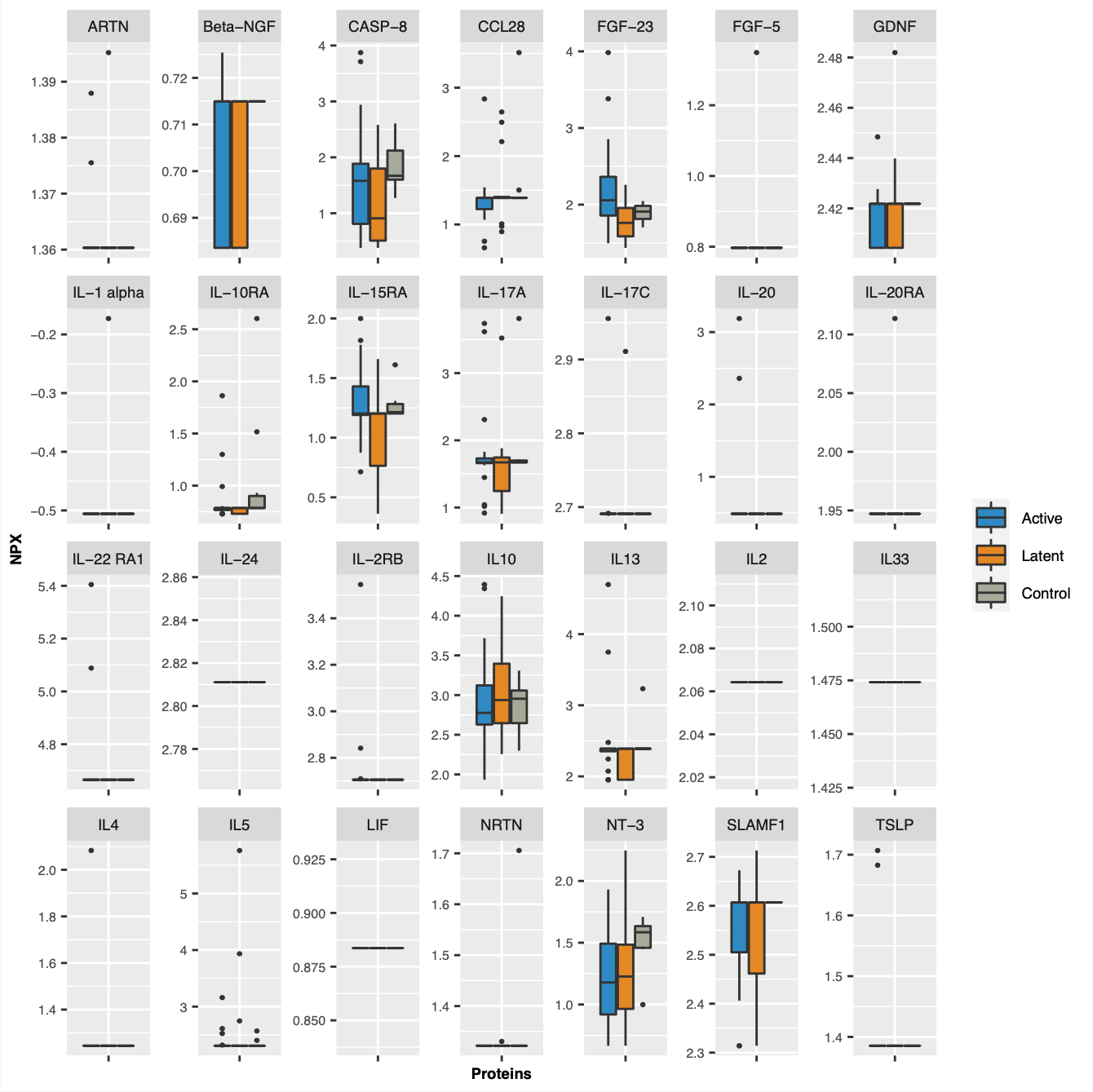

Figure 3. The NPX values of removed proteins in different groups of individuals.

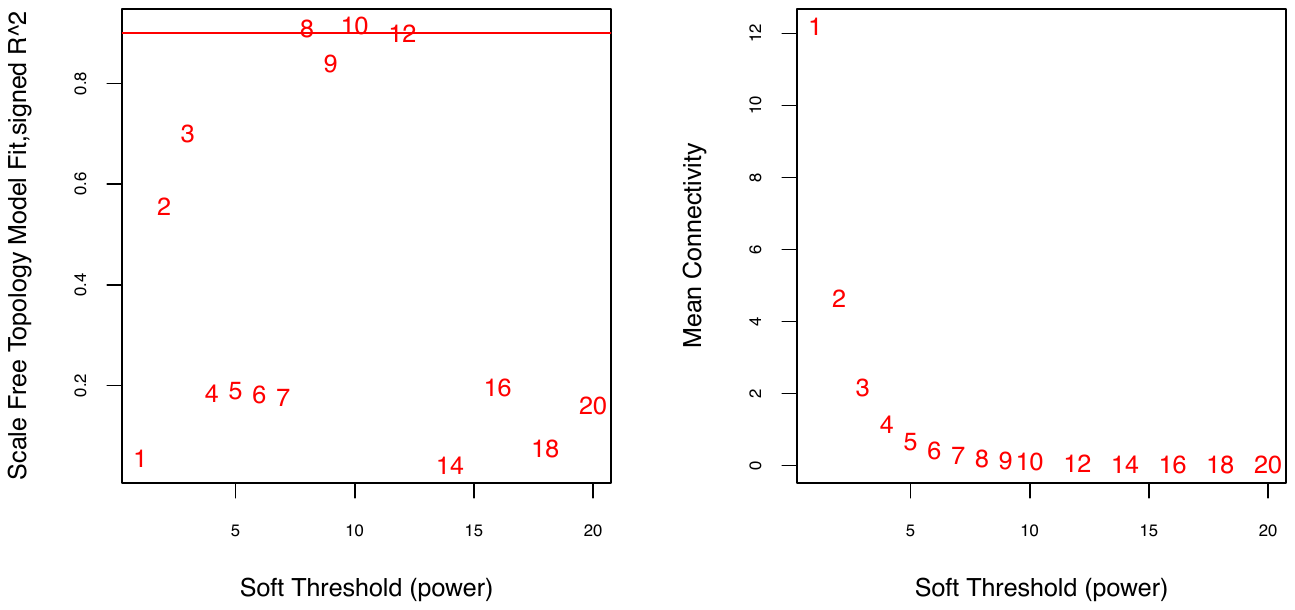

Figure 4. The power parameter for the Scale-free topology.

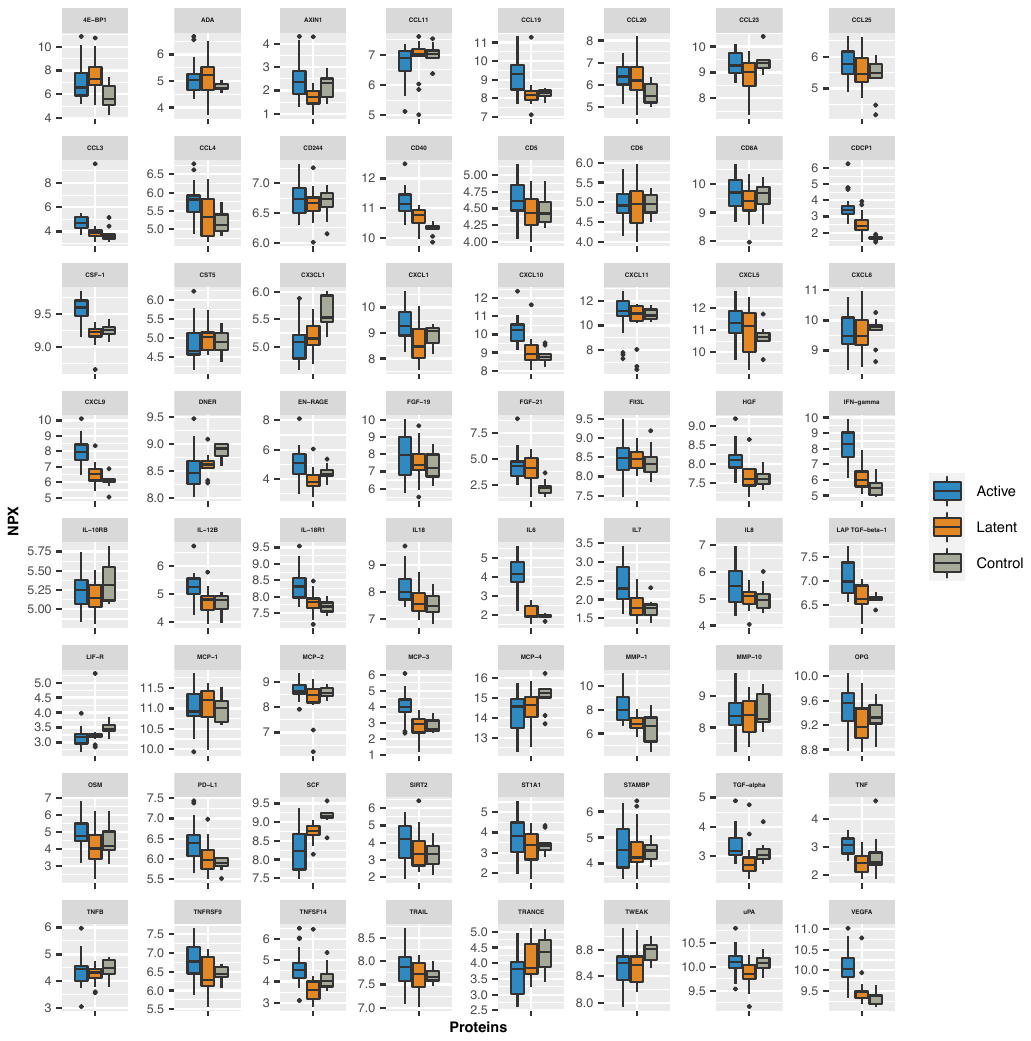

Figure 5. The NPX values of all remaining proteins in different groups of individuals.

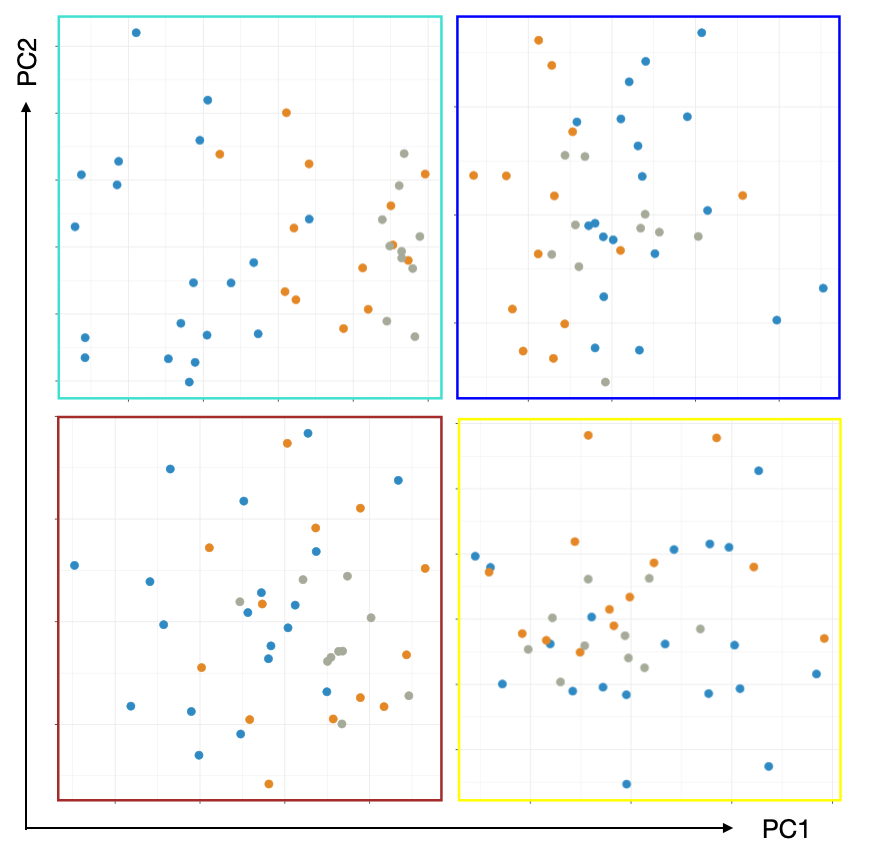

Figure 6. Visualizing samples regarding PC1 and PC2, obtained from the expression data of each module. Modules are surrounded indicated by colours including turquoise, blue, brown and yellow. Each point in plots represents one sample from one out of three groups of active TB (blue), latent TB (orange) and healthy control (grey).

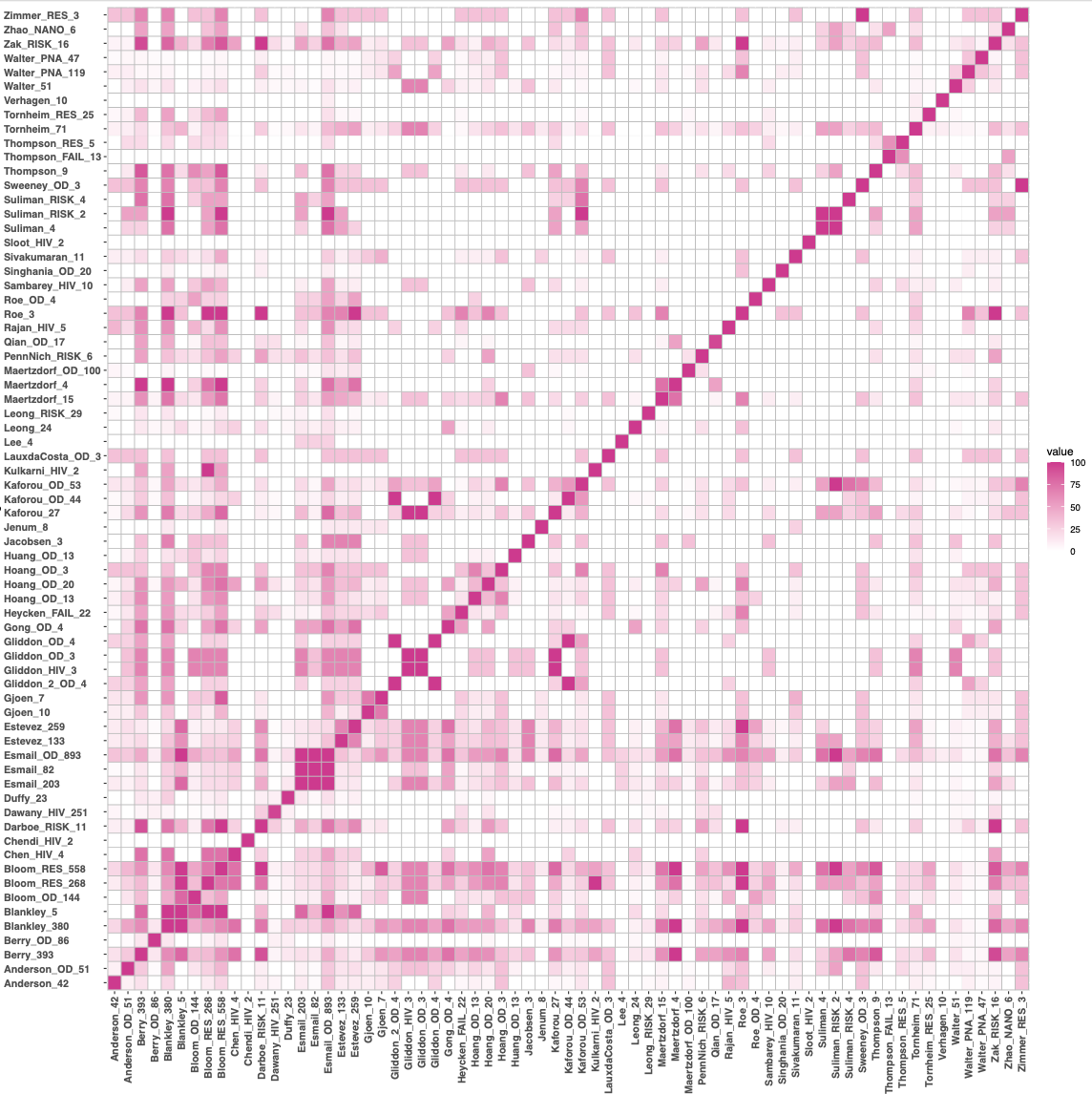

Figure 7. The overlap between the other published gene signatures from the TBSignatureProfiler R package. The value in each cell matrix indicates the percentage of genes that are shared between two distinct gene signatures.

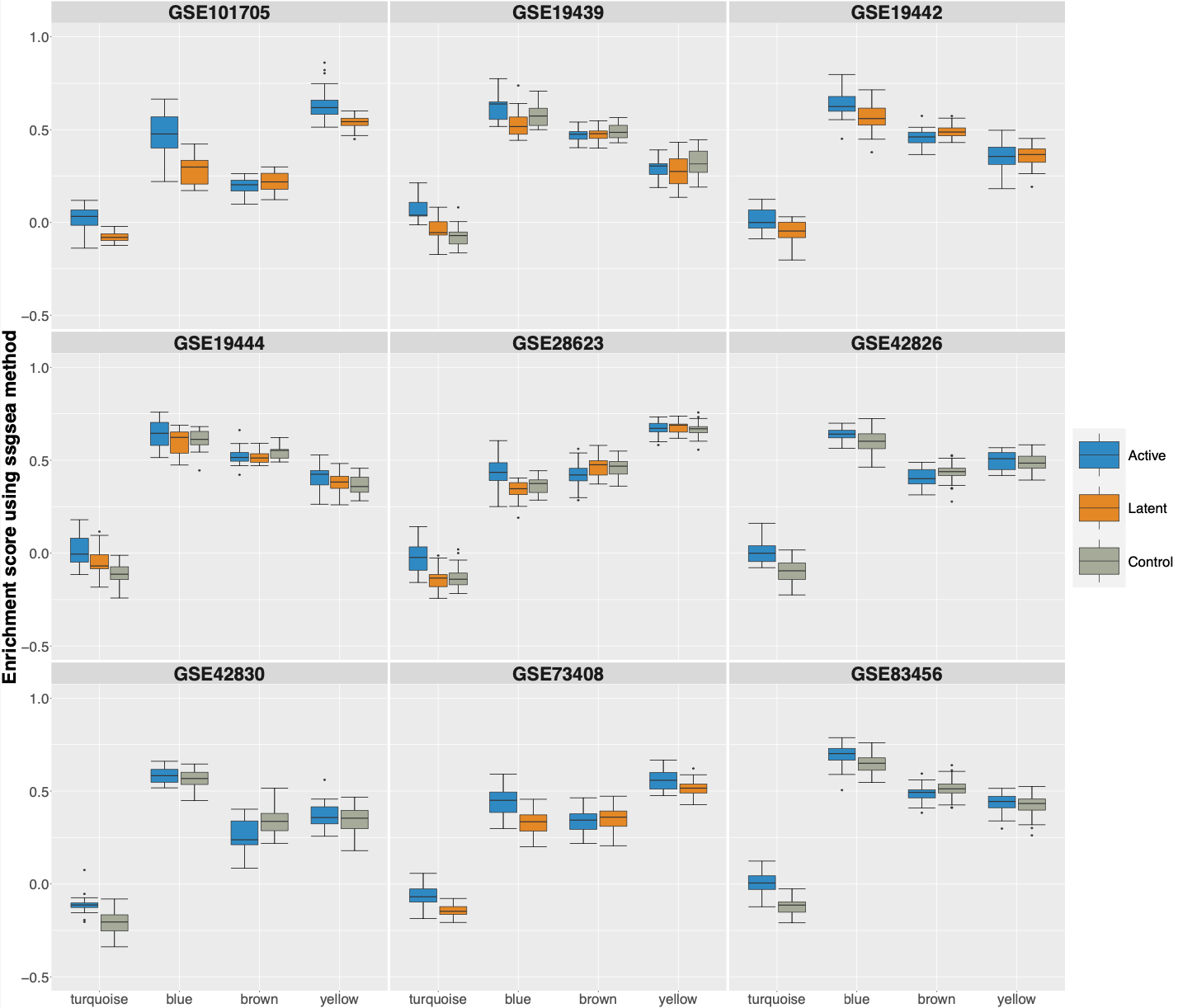

Figure 8. The enrichment analysis of all modules (from left to right: turquoise, blue, brown and yellow) on different transcriptomic datasets using the ssgsea method.

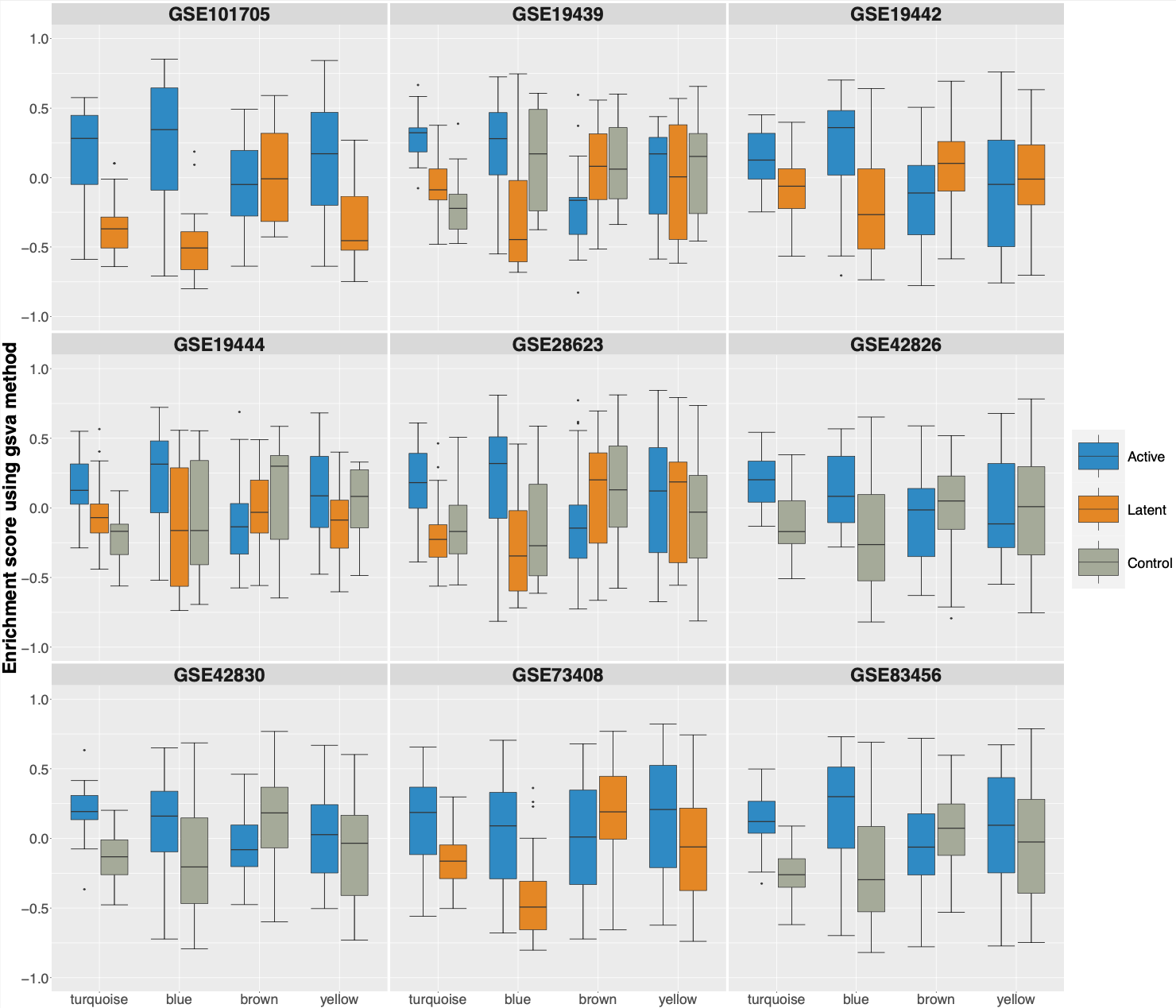

Figure 9. The enrichment analysis of all modules (from left to right: turquoise, blue, brown and yellow) on different transcriptomic datasets using the gsva method.

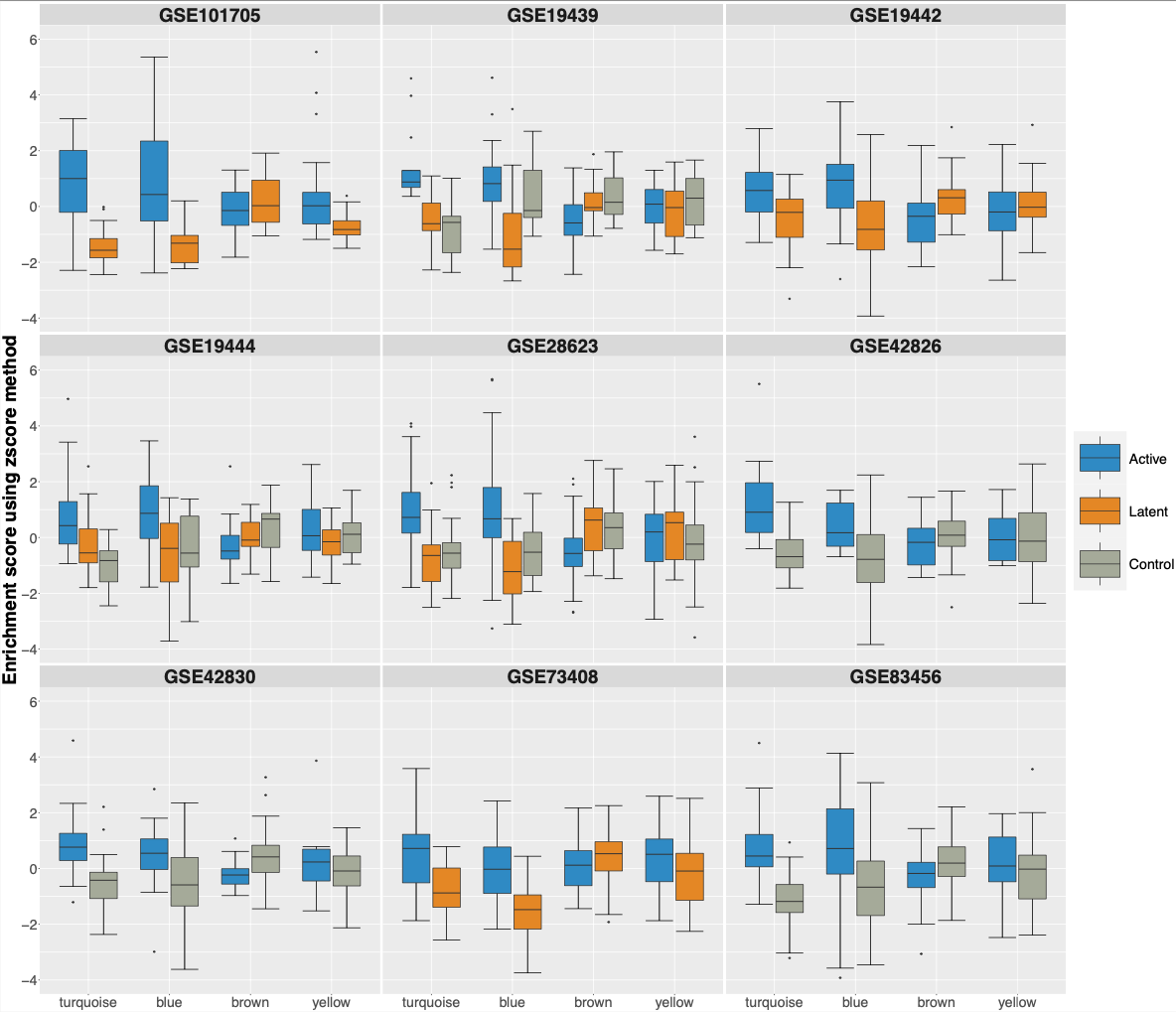

Figure 10. The enrichment analysis of all modules (from left to right: turquoise, blue, brown and yellow) on different transcriptomic datasets using the zscore method.

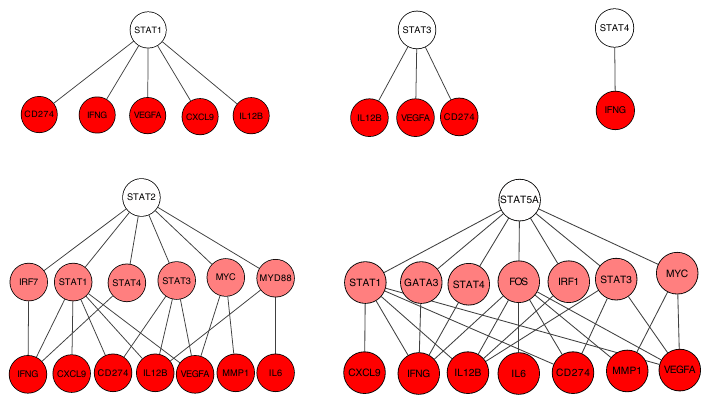

Figure 11. The signalling pathways between different signal transducer and activator of transcription (STAT) transcription factors and the proteins of our signature regarding the KEGG database.

Table 1. Details of Active TB patients

| **Active TB** | n=19 |  |
| --- | --- | --- |
| **Symptoms** |  |  |
| cough, n | 11 |  |
| fever/night sweats, n | 12 |  |
| duration, mo (mean, range) | 4,5 (1-24) |  |
| **Manifestation** |  |  |
| pulmonary/pleuritis (+/-lgll) | 15 |  |
| lymphnode | 1 |  |
| disseminated | 2 |  |
| Other* | 1 |  |
| **CXR/CT** |  |  |
| any pathology | 16 |  |
| infiltrates | 15 |  |
| cavities | 14 |  |
| pleural effusion | 3 |  |
| **Mycobacteriology** |  |  |
| sputum microscopy pos | 5 |  |
| any sample microscopy pos | 6 |  |
| PCR positive | 11 |  |
| culture positive | 18 |  |
| **Biochemistry** | Mean (range) | n outside the reference interval |
| CRP | 25 (1-94) | 13 (>10) |
| ESR | 51 (7-117) | 14 (>20) |
| leukocytes | 6.5 (3.8-11.8) | 2 (>9.5) |
| hemoglobine | 126 (102-149) | 8 (F<120, M<130) |
| albumine (n=18) | 32 (26-38) | 15 (<38) |

*other= soft tissue abscess

Table 2. Plasma proteins analyzed by inflammation panel of Olink

| Protein Name (Short Name) | Uniprot ID |
| --- | --- |
| Adenosine Deaminase (ADA) | P00813 |
| Artemin (ARTN) | Q5T4W7 |
| Axin-1 (AXIN1) | O15169 |
| Beta-nerve growth factor (Beta-NGF) | P01138 |
| Brain-derived neurotrophic factor (BDNF) | P23560 |
| Caspase-8 (CASP-8 ) | Q14790 |
| C-C motif chemokine 3 (CCL3) | P10147 |
| C-C motif chemokine 4 (CCL4 ) | P13236 |
| C-C motif chemokine 19 (CCL19) | Q99731 |
| C-C motif chemokine 20 (CCL20) | P78556 |
| C-C motif chemokine 23 (CCL23) | P55773 |
| C-C motif chemokine 25 (CCL25) | O15444 |
| C-C motif chemokine 28 (CCL28) | Q9NRJ3 |
| CD40L receptor (CD40) | P25942 |
| CUB domain-containing protein 1 (CDCP1) | Q9H5V8 |
| C-X-C motif chemokine 1 (CXCL1) | P09341 |
| C-X-C motif chemokine 5 (CXCL5 ) | P42830 |
| C-X-C motif chemokine 6 (CXCL6) | P80162 |
| C-X-C motif chemokine 9 (CXCL9 ) | Q07325 |
| C-X-C motif chemokine 10 (CXCL10 ) | P02778 |
| C-X-C motif chemokine 11 (CXCL11) | O14625 |
| Cystatin D (CST5) | P28325 |
| Delta and Notch-like epidermal growth factor-related receptor (DNER) | Q8NFT8 |
| Eotaxin (CCL11) | P51671 |
| Eukaryotic translation initiation factor 4E-binding protein 1 (4E-BP1) | Q13541 |
| Fibroblast growth factor 21 (FGF-21) | Q9NSA1 |
| Fibroblast growth factor 23 (FGF-23) | Q9GZV9 |
| Fibroblast growth factor 5 (FGF-5) | Q8NF90 |
| Fibroblast growth factor 19 (FGF-19) | O95750 |
| Fms-related tyrosine kinase 3 ligand (Flt3L) | P49771 |
| Fractalkine (CX3CL1 ) | P78423 |
| Glial cell line-derived neurotrophic factor (GDNF) | P39905 |
| Hepatocyte growth factor (HGF) | P14210 |
| Interferon gamma (IFN-gamma) | P01579 |
| Interleukin-1 alpha (IL-1 alpha) | P01583 |
| Interleukin-2 (IL-2) | P60568 |
| Interleukin-2 receptor subunit beta (IL-2RB) | P14784 |
| Interleukin-4 (IL-4) | P05112 |
| Interleukin-5 (IL5) | P05113 |
| Interleukin-6 (IL6) | P05231 |
| Interleukin-7 (IL-7) | P13232 |
| Interleukin-8 (IL-8) | P10145 |
| Interleukin-10 (IL10) | P22301 |
| Interleukin-10 receptor subunit alpha (IL-10RA) | Q13651 |
| Interleukin-10 receptor subunit beta (IL-10RB) | Q08334 |
| Interleukin-12 subunit beta (IL-12B) | P29460 |
| Interleukin-13 (IL-13) | P35225 |
| Interleukin-15 receptor subunit alpha (IL-15RA) | Q13261 |
| Interleukin-17A (IL-17A) | Q16552 |
| Interleukin-17C (IL-17C) | Q9P0M4 |
| Interleukin-18 (IL-18) | Q14116 |
| Interleukin-18 receptor 1 (IL-18R1) | Q13478 |
| Interleukin-20 (IL-20) | Q9NYY1 |
| Interleukin-20 receptor subunit alpha (IL-20RA) | Q9UHF4 |
| Interleukin-22 receptor subunit alpha-1 (IL-22 RA1) | Q8N6P7 |
| Interleukin-24 (IL-24) | Q13007 |
| Interleukin-33 (IL-33) | O95760 |
| Latency-associated peptide transforming growth factor beta-1 (LAP TGF-beta-1) | P01137 |
| Leukemia inhibitory factor (LIF) | P15018 |
| Leukemia inhibitory factor receptor (LIF-R) | P42702 |
| Macrophage colony-stimulating factor 1 (CSF-1) | P09603 |
| Matrix metalloproteinase-1 (MMP-1) | P03956 |
| Matrix metalloproteinase-10 (MMP-10) | P09238 |
| Monocyte chemotactic protein 1 (MCP-1) | P13500 |
| Monocyte chemotactic protein 2 (MCP-2) | P80075 |
| Monocyte chemotactic protein 3 (MCP-3) | P80098 |
| Monocyte chemotactic protein 4 (MCP-4) | Q99616 |
| Natural killer cell receptor 2B4 (CD244) | Q9BZW8 |
| Neurotrophin-3 (NT-3) | P20783 |
| Neurturin (NRTN) | Q99748 |
| Oncostatin-M (OSM) | P13725 |
| Osteoprotegerin (OPG) | O00300 |
| Programmed cell death 1 ligand 1 (PD-L1) | Q9NZQ7 |
| Protein S100-A12 (EN-RAGE ) | P80511 |
| Signaling lymphocytic activation molecule (SLAMF1) | Q13291 |
| SIR2-like protein 2 (SIRT2) | Q8IXJ6 |
| STAM-binding protein (STAMBP) | O95630 |
| Stem cell factor (SCF) | P21583 |
| Sulfotransferase 1A1 (ST1A1) | P50225 |
| T cell surface glycoprotein CD6 isoform (CD6) | Q8WWJ7 |
| T-cell surface glycoprotein CD5 (CD5) | P06127 |
| Thymic stromal lymphopoietin (TSLP) | Q969D9 |
| TNF-beta (TNFB) | P01374 |
| TNF-related activation-induced cytokine (TRANCE) | O14788 |
| TNF-related apoptosis-inducing ligand (TRAIL) | P50591 |
| Transforming growth factor alpha (TGF-alpha) | P01135 |
| Tumor necrosis factor (Ligand) superfamily, member 12 (TWEAK) | O43508 |
| Tumor necrosis factor (TNF) | P01375 |
| Tumor necrosis factor ligand superfamily member 14 (TNFSF14 ) | O43557 |
| Tumor necrosis factor receptor superfamily member 9 (TNFRSF9) | Q07011 |
| Urokinase-type plasminogen activator (uPA) | P00749 |
| Vascular endothelial growth factor A (VEGF-A) | P15692 |

Table 3. More details about selected transcriptomic TB datasets from curatedTBData R package

| **Dataset** | **GeographicalRegion** | **Tissue** | **Age** | **HIVStatus** | **Control** | **Latent** | **Active** |
| --- | --- | --- | --- | --- | --- | --- | --- |
| **GSE73408** | US | Whole Blood | >18 | Negative | NA | 35 | 35 |
| **GSE101705** | South India | Whole Blood | >18 | Negative | NA | 16 | 28 |
| **GSE19439** | UK | Whole Blood | 19-72 | Negative | 12 | 17 | 13 |
| **GSE19442** | South Africa | Whole Blood | 18-48 | Negative | NA | 31 | 20 |
| **GSE19444** | UK | Whole Blood | >18 | Negative | 12 | 21 | 21 |
| **GSE42826** | Germany | Whole Blood | >17 | Negative | 52 | NA | 11 |
| **GSE42830** | Germany | Whole Blood | >17 | Negative | 38 | NA | 16 |
| **GSE83456** | UK | Whole Blood | NA | Negative | 61 | NA | 45 |
| **GSE28623** | The Gambia | Whole Blood | 16-53 | Negative | 37 | 25 | 46 |
